# Ethnic Discrimination is Related to Increased Substance Use and Intentions to Use Across Diverse Groups of Adolescents

**DOI:** 10.1101/2024.10.02.24314798

**Authors:** Robert Rosales, Philip Veliz, John Jardine, Alexander Weigard, Sean Esteban McCabe

## Abstract

**Introduction:** In recent years, adolescents of color report greater use of selected substances than white adolescents, including alcohol, tobacco, and cannabis. Increased levels of discrimination during the COVID-19 pandemic may have added to the chronic burden associated with increased substance use among adolescents of color. This study assessed the prevalence of substance use (alcohol, tobacco and cannabis) and intentions to use among adolescents by race/ethnicity and assessed associations between discrimination and substance use outcomes across groups.

**Methods:** The data come from the national panel of 11,868 adolescents in the Adolescent Brain Cognitive Development study (baseline through 4^th^ follow-up). We tested the prevalence rates of substance use and intentions by race/ethnicity. Multivariable longitudinal analyses tested whether 1) discrimination was connected to substance use and intentions, and 2) whether that relationship differed by race/ethnicity.

**Results:** White adolescents reported the greatest use and curiosity about alcohol. Black adolescents reported the highest rates of being willing to try any of the substances. Hispanic adolescents reported the highest rates of cannabis use. Multiracial adolescents reported the highest rates of tobacco use, curiosity about tobacco, and curiosity about cannabis. Discrimination was found to be associated with greater substance use among all racial/ethnic groups, except Black adolescents for alcohol use and Asian adolescents for alcohol use and tobacco use.

**Conclusion:** This study supports national trends about substance use disparities among adolescents of color. Findings from this study also show that discrimination may explain some of these increased trends through intentions to use substances.

---

Substance use disorders (SUDs) cause a significant social and economic burden worldwide; in the United States (U.S.) alone more than 65 million U.S. adults will have an alcohol use disorder in their lifetime, 8.5% of adults report a nicotine dependance in the past month, 6.8% reported a cannabis use disorder in the past year, and 9.6% of adults report any drug use disorder in the past year (SAMHSA, 2022, 2024). In past years, the major burden of SUDs has been concentrated mostly among white people in the U.S. (Miech et al., 2024; Miech et al., 2023). However, Black and Hispanic adolescents in the 2022 and 2023 Monitoring the Future (MTF) study reported that they are more likely to use most substances (e.g., alcohol, cannabis, and all forms of tobacco) than white adolescents, a reversal from years prior to 2022 (Miech et al., 2024; Miech et al., 2023). This difference in substance use rates may signal future racial differences in SUD as adolescents age into adulthood.

The Lifespan Biopsychosocial Model of Cumulative Vulnerability and Minority Health posits that discrimination is related to substance use disparities due to the chronic stress burden that accumulates over time to diminish a person’s ability to cope without relying on substances (Myers, 2009). It is possible that people of color’s heightened stress during the COVID-19 pandemic and increasing racism may partially explain the increase in substance use (Leventhal et al., 2018; Tao et al., 2023). Specifically, adolescents of color may have used substances at greater rates in recent years because of the chronic discrimination they experienced in earlier childhood that has accumulated with daily ethnic discrimination.

The latest MTF data shows that differences in substance use between adolescents of color and white adolescents start in 10^th^ grade (Miech et al., 2024; Miech et al., 2023). However, according to the Theory of Planned behavior, attitudes (e.g., curiosity about using) and intentions to use (e.g., willing to try soon) can predict future planned and intentional substance use behavior (Ajzen, 1991; Jackson et al., 2014; Marcoux and Shope, 1997). Willingness and curiosity to try substances have been shown to predict future use, including among Hispanic adolescents (Kam et al., 2009; Maher and Rickwood, 1998; Malmberg et al., 2012; Nodora et al., 2014; Pierce et al., 2005). Hence, substance use risk can be assessed prior to 10^th^ grade by measuring these key precursors. Little is known about the relationship between discrimination and intentions to use (Wade et al., 2021), especially among adolescents of color.

## Present study

One of the largest publicly available datasets of adolescent health (Adolescent Brain Cognitive Development [ABCD]) was used to assess: 1) the prevalence of substance use, 2) whether ethnic discrimination is related to substance use and intentions to use, and 3) whether this relationship differed by race/ethnicity.

## Methods

We conducted secondary data analyses of the ABCD data, which has collected repeated measures responses from nearly 11,868 adolescents recruited nationally over 10 years. We used the publicly available panel data from baseline to 4^th^ follow-up (2017 to present), which includes a sample of 1,784 non-Hispanic Black (15%; hereinafter Black), 2,410 Hispanic (20.3%), 252 non-Hispanic Asian (2.1%; hereinafter Asian), 1,247 non-Hispanic other race or multiracial (10.5%; hereinafter other race/multiracial), and 6,173 non-Hispanic white (52.1%; hereinafter white). A greater percent of the adolescent participants identified as male (52.2%). In terms of family makeup, the largest proportion of the participants’ caregivers had a bachelor’s degree or higher (59.5%), married (67.8%), worked full-time (84.8%), and made $100,000 or greater total combined family income (38.4%).

### Measures

#### Ethnic discrimination

Perceived Ethnic Discrimination (PED) Scale is a 7-item measure created to assess perceptions of ethnic discrimination among minority and immigrant adolescents using items like, “Others behave in an unfair or negative way toward my ethnic group” and “I don’t feel accepted by other Americans” (Phinney et al., 1998). The PED uses a 5 point scale ranging from 0 (almost never) to 4 (very often) and has shown high reliability in past research (e.g., α=81-90; (Cano et al., 2015; Phinney et al., 1998). Responses are averaged with higher responses indicating greater perceived ethnic discrimination. We used respondents’ highest scores across the year 1, 2, and 4 follow-ups.

#### Substance use and intentions to use

Substance use was evaluated at baseline (lifetime use) and each follow-up event (use since the last session was completed) using the “substance use interview” and “substance use phone interview (mid-year)” modules. For alcohol, “use” is defined as having tried a full drink of beer, wine, or liquor. For tobacco and marijuana, “use” is defined as having tried any tobacco/nicotine or marijuana product, including just a puff. More details are given in the table footnotes.

In terms of intentions to use, the curiosity items were measured with three questions: “Have you ever been curious about…” 1) “using a tobacco product such as cigarettes, e-cigarettes, hookah, or cigars?”, 2) “drinking alcohol?”, and 3) “trying marijuana?” These measures were dichotomized such that 0 = “not at all curious” and 1 = “a little curious”, “somewhat curious”, or “very curious”. The willingness to try items were also measured with three questions: “Do you think you will try…” 1) “a tobacco product soon?”, 2) “alcohol soon?”, and 3) “marijuana soon?” These measures were dichotomized such that 0 = “definitely not” or “probably not” and 1 = “definitely yes” or “probably yes”.

#### Race/ethnicity

Race/ethnicity was measured with two items that asked the parent, “What race do you consider the child to be?” and “Do you consider the child Hispanic/Latino/Latina?” The ABCD recoded these responses into a single, five-category variable: 1) white, 2) Black, 3) Hispanic, 4) Asian, and 5) other race or multiracial.

### Analytic plan

All data analyses were conducted in Stata 18.0, statistical software. Bivariate analyses were conducted to assess the prevalence of substance use and intentions to use substances by race/ethnicity and by level of discrimination. Generalized estimating equations (with an exchangeable correlation matrix) were used to assess the longitudinal association between race/ethnicity, discrimination and 1) substance use, and 2) intentions to use. We then conducted interaction effects models to assess how the level of discrimination was associated with substance use/intentions to use by different racial/ethnic groups. Each model was conducted separately to assess the outcome, which included nine models for main effects (models 1-9) and nine for interaction effects only (models 10-18). All outcomes were time-varying; all covariates (except time) were time-invariant. All models are adjusted for time, the respondent’s age at baseline, the respondent’s sex, parental marital status, parental education level, parental employment status, and total combined family income.

## Results

Table 1 presents the prevalence rates of substance use and intentions by race/ethnicity. White adolescents reported the highest rates of alcohol use and curiosity about alcohol. Black adolescents reported the highest rates of being willing to try alcohol, tobacco, and marijuana. Hispanic adolescents reported the highest rates of marijuana use. Other race or multiracial adolescents reported the highest rates of tobacco use, curiosity about tobacco, and curiosity about marijuana.

**Table 1:** Prevalence of Substance Use and Intentions to Use by Race / Ethnicity and Discrimination: % (95% CI)

| Race / Ethnicity | Substance Use |  |  | Intentions to Use |  |  |  |  |  |
| --- | --- | --- | --- | --- | --- | --- | --- | --- | --- |
|  | Alcohol | Tobacco | Cannabis | Curiosity |  |  | Will Try Soon |  |  |
|  |  |  |  | Alcohol | Tobacco | Cannabis | Alcohol | Tobacco | Cannabis |
| Non-Hispanic White only | 3.76 (3.30, 4.28) | 5.28 (4.72, 5.91) | 2.80 (2.41, 3.25) | 36.13 (34.69, 37.60) | 30.03 (28.85, 31.24) | 15.10 (14.18, 16.07) | 2.73 (2.29, 3.25) | 1.52 (1.24, 1.87) | 1.17 (0.93, 1.48) |
| Non-Hispanic Black only | 1.12 (0.72, 1.73) | 4.76 (3.87, 5.85) | 3.48 (2.71, 4.45) | 26.46 (24.23, 28.81) | 25.62 (23.50, 27.87) | 15.31 (13.61, 17.19) | 6.13 (5.03, 7.44) | 3.65 (2.86, 4.64) | 3.47 (2.69, 4.47) |
| Hispanic | 2.78 (2.17, 3.55) | 6.02 (5.08, 7.11) | 3.90 (3.18, 4.79) | 35.11 (32.97, 37.31) | 30.78 (28.91, 32.72) | 14.67 (13.25, 16.21) | 5.73 (4.75, 6.89) | 2.76 (2.15, 3.52) | 2.10 (1.57, 2.79) |
| Non-Hispanic Asian only | 0.40 (0.06, 2.79) | 0.79 (0.20, 3.14) | 0.79 (0.20, 3.14) | 32.47 (26.31, 39.31) | 27.76 (22.37, 33.87) | 10.68 (7.10, 15.76) | 4.62 (2.40, 8.68) | 0.40 (0.06, 2.84) | 0.43 (0.06, 3.02) |
| Non-Hispanic other race or multiracial | 3.05 (2.21, 4.19) | 7.54 (6.16, 9.19) | 3.53 (2.62, 4.74) | 32.58 (29.59, 35.72) | 32.16 (29.54, 34.89) | 18.26 (16.13, 20.60) | 4.11 (2.99, 5.63) | 2.95 (2.14, 4.06) | 2.16 (1.47, 3.15) |

**Notes.** For calculating prevalence of substance use (and intentions to use), respondents were coded as ‘1’ if they indicated use at any event. Else, they were coded as ‘0’ as long as they indicated no use at at least one event. For the models in table 2, substance use and intentions to use were left as time-varying. For alcohol, ‘use’ is defined as having tried a full drink of beer, wine, or liquor. For tobacco, ‘use’ is defined as having tried a tobacco cigarette; an electronic cigarette, vape pen, or e-hookah; smokeless tobacco, chew, or snus; cigars, including traditional cigars, little cigars, or cigarillos; hookah; pipes; and nicotine replacements, such as patches, gums, nasal sprays, inhalers, and lozenges. A puff of a tobacco product counts as ‘use’. For marijuana, ‘use’ is defined as having tried smoking marijuana; blunts; marijuana that you eat, such as pot cookies, gummy bears, or brownies; marijuana oils or concentrates; marijuana infused alcohol drinks; concentrated marijuana tinctures; vaping marijuana flower or bud; and vaping marijuana oils or concentrates. More detailed examples of certain methods are given in the ABCD codebooks. A puff of a marijuana product counts as ‘use’.
For a given event, only respondents who knew what the substance was, but hadn’t tried it yet (including just a sip of alcohol or a puff of tobacco / marijuana), were asked about their curiosity and willingness to try that substance.

Table 2 presents the main effects of discrimination on substance use and intentions. Across all three substances, greater discrimination was robustly associated with greater odds of use (aOR = 1.86 – 1.92), curiosity (aOR = 1.37 – 1.68), and willingness to use (aOR = 1.62 – 2.08)

**Table 2:** Adjusted Odds Ratios for Substance Use and Intentions to Use as Functions of Race / Ethnicity and Discrimination: aOR (95% CI)

|  | Substance Use |  |  | Intentions to Use |  |  |  |  |  |
| --- | --- | --- | --- | --- | --- | --- | --- | --- | --- |
|  |  |  |  | Curiosity |  |  | Will Try Soon |  |  |
|  | Alcohol<br>n = 11,257<br>obs. =<br>47,834 | Tobacco<br>n = 11,257<br>obs. =<br>47,834 | Cannabis<br>n = 11,257<br>obs. =<br>47,831 | Alcohol<br>n = 8,657<br>obs. =<br>32,474 | Tobacco<br>n = 11,122<br>obs. =<br>40,713 | Cannabis<br>n = 10,905<br>obs. =<br>32,714 | Alcohol<br>n = 8,660<br>obs. =<br>32,421 | Tobacco<br>n = 11,121<br>obs. =<br>41,005 | Cannabis<br>n = 10,905<br>obs. =<br>32,794 |
| **Main Effects:**<br>**Models 1-9** |  |  |  |  |  |  |  |  |  |
| Non-Hispanic White only | Ref. | Ref. | Ref. | Ref. | Ref. | Ref. | Ref. | Ref. | Ref. |
| Non-Hispanic Black only | 0.26 (0.15,<br>0.43) | 0.44 (0.33,<br>0.58) | 0.70 (0.50,<br>0.98) | 0.76 (0.66,<br>0.88) | 0.74 (0.65,<br>0.85) | 0.81 (0.67,<br>0.97) | 1.83 (1.29,<br>2.59) | 1.19 (0.80,<br>1.76) | 1.59 (1.04,<br>2.43) |
| Hispanic | 0.76 (0.56,<br>1.02) | 0.78 (0.63,<br>0.98) | 0.95 (0.72,<br>1.26) | 1.12 (1.00,<br>1.25) | 0.97 (0.87,<br>1.08) | 0.84 (0.72,<br>0.98) | 2.03 (1.50,<br>2.75) | 1.11 (0.77,<br>1.61) | 1.05 (0.69,<br>1.59) |
| Non-Hispanic Asian only | 0.08 (0.01,<br>0.66) | 0.11 (0.02,<br>0.58) | 0.26 (0.06,<br>1.19) | 0.81 (0.61,<br>1.07) | 0.92 (0.70,<br>1.19) | 0.62 (0.41,<br>0.94) | 2.18 (1.11,<br>4.28) | 0.30 (0.04,<br>2.16) | 0.37 (0.05,<br>2.68) |
| Non-Hispanic other race or<br>multiracial | 0.69 (0.48,<br>0.99) | 1.01 (0.79,<br>1.30) | 0.94 (0.66,<br>1.33) | 0.98 (0.85,<br>1.12) | 1.03 (0.91,<br>1.17) | 1.21 (1.02,<br>1.43) | 1.46 (0.99,<br>2.14) | 1.37 (0.90,<br>2.09) | 1.41 (0.88,<br>2.26) |
| Discrimination score | 1.92 (1.63,<br>2.27) | 1.88 (1.67,<br>2.11) | 1.86 (1.61,<br>2.15) | 1.37 (1.27,<br>1.48) | 1.61 (1.50,<br>1.72) | 1.68 (1.53,<br>1.83) | 1.62 (1.38,<br>1.90) | 1.77 (1.48,<br>2.11) | 2.08 (1.74,<br>2.49) |
| **Interaction Effects:**<br>**Models 10-18** |  |  |  |  |  |  |  |  |  |
| Non-Hispanic White only x<br>Discrimination | 2.48 (2.00,<br>3.06) | 2.32 (1.96,<br>2.75) | 2.33 (1.88,<br>2.90) | 1.34 (1.18,<br>1.53) | 1.71 (1.52,<br>1.91) | 1.61 (1.39,<br>1.88) | 1.27 (0.88,<br>1.84) | 1.87 (1.38,<br>2.53) | 1.72 (1.17,<br>2.53) |
| Non-Hispanic Black only x<br>Discrimination | 0.91 (0.59,<br>1.40) | 1.22 (0.98,<br>1.52) | 1.38 (1.07,<br>1.77) | 1.21 (1.07,<br>1.36) | 1.36 (1.21,<br>1.52) | 1.57 (1.37,<br>1.80) | 1.66 (1.33,<br>2.07) | 1.75 (1.38,<br>2.22) | 2.58 (2.10,<br>3.17) |
| Hispanic only x Discrimination | 1.63 (1.21,<br>2.20) | 1.82 (1.51,<br>2.21) | 1.79 (1.42,<br>2.27) | 1.42 (1.24,<br>1.61) | 1.50 (1.33,<br>1.70) | 1.55 (1.32,<br>1.81) | 1.77 (1.37,<br>2.29) | 1.63 (1.19,<br>2.22) | 1.77 (1.26,<br>2.50) |
| Non-Hispanic Asian only x<br>Discrimination | 0.85 (0.21,<br>3.54) | 0.63 (0.13,<br>2.98) | 1.35 (0.36,<br>5.01) | 1.20 (0.73,<br>1.97) | 1.82 (1.21,<br>2.73) | 1.37 (0.77,<br>2.42) | 5.93 (3.13,<br>11.21) | 3.01 (0.98,<br>9.26) | 3.22 (1.04,<br>9.92) |
| Non-Hispanic other race or<br>multiracial only x Discrimination | 1.60 (1.17,<br>2.19) | 1.91 (1.54,<br>2.38) | 1.93 (1.47,<br>2.54) | 1.49 (1.28,<br>1.74) | 1.80 (1.57,<br>2.06) | 2.02 (1.71,<br>2.39) | 1.93 (1.43,<br>2.62) | 2.04 (1.47,<br>2.83) | 2.01 (1.36,<br>2.97) |

**Notes.** All models are adjusted for time, the respondent's age at baseline, the respondent's sex, parental marital status, parental education level, parental employment status, and total combined family income. All outcomes are time-varying; all covariates (except time) are time-invariant. Main effects for race / ethnicity and discrimination were not included in models 10-18.

Table 2 also presents the interaction effects of race/ethnicity on the relationship between discrimination and substance use and intentions. Greater discrimination was consistently associated with greater odds of use, curiosity, and willingness to use substances among all race/ethnic groups in this study, except for Black adolescents reporting lower odds of alcohol use and Asian adolescents reporting lower odds of alcohol and tobacco use when experiencing discrimination. The aORs for this interaction term were uniformly positive and substantial across all groups for both curiosity (aOR = 1.20 – 2.02) and willingness (aOR = 1.27 – 5.93) to use.

## Discussion

Due to the high prevalence of substance use among racial/ethnic minorities in recent years, this study assessed the prevalence of substance use and intentions as well as the association between discrimination experiences and substance use among racial/ethnic adolescent groups. The findings from the present study reinforced some of the results from other national studies showing higher substance use among Hispanic and Black adolescents when compared with white adolescents (Miech et al., 2024; Miech et al., 2023). The present study found that Black, Hispanic, and other/multiracial adolescents reported greater tobacco and cannabis use than white adolescents. Multiracial adolescents had the highest prevalence of substance use, consistent with a growing body of research (Goings et al., 2018; Hai et al., 2024; Jackson and Lecroy, 2009). Although white adolescents reported greater prevalence of alcohol use, they reported the lowest willingness to try alcohol soon when compared with Black, Hispanic, Asian, and other/multiracial adolescents. Most of the ABCD sample in the latest publicly available wave have not reached 10^th^ grade yet, suggesting that the majority of alcohol use initiation will occur at subsequent timepoints. Thus, Black, Hispanic, and other/multiracial adolescents increased willingness to try alcohol may point to greater future alcohol initiation in these groups, leading to future disparities in alcohol use. Startingly, the prevalence of willingness to use tobacco and cannabis were also higher among Black, Hispanic, and other/multiracial adolescents when compared to white adolescents. Prior research has shown that adolescents of color who report being more willing to try alcohol, tobacco, and cannabis are more likely to use those substances in the future (Kam et al., 2009). Thus, willingness to use findings in this study likely point to future increased disparities in tobacco and cannabis use for Black, Hispanic, and other/multiracial adolescents when compared to their white counterparts.

Other race/multiracial adolescents reported the greatest prevalence of curiosity towards tobacco and cannabis when compared to all other racial/ethnic groups in this study. It is possible that the high risk we see in research related to substance use among multiracial adolescents may be related to the greater prevalence of curiosity towards these substances (Goings et al., 2018; Hai et al., 2024; Jackson and Lecroy, 2009). Meanwhile, white adolescents reported the greatest curiosity towards alcohol use, which may explain the current higher prevalence of alcohol among white adolescents.

The findings related to discrimination support models suggesting that discrimination is associated with increased substance use, curiosity, and willingness to use (Myers, 2009). Although there is a plethora of research showing the connection between discrimination and substance use among adolescents of color (Brody et al., 2012; Dai et al., 2024; Gerrard et al., 2012; Gibbons et al., 2010; Gibbons et al., 2004; Myers, 2009), our findings show white adolescents reported the greatest odds of substance use when discrimination scores increased. However, they reported lower odds of willing to try alcohol, tobacco, and cannabis than most of the adolescents of color groups. This finding signal that substance use differences may widen. If this finding translates to future use, the effects of discrimination on substance use may be stronger as adolescents get to 10^th^ grade and later.

## Limitations

This study is not without limitation. First, we combined adolescents from various racial/ethnic groups that may be heterogeneous into five groups. Future studies should assess whether country of origin, skin color, and other differences within racial/ethnic groups may be related to variance in these relationships. Further, substance use is still relatively low in this young cohort of adolescents making it difficult to assess differences across racial/ethnic groups due to sample size constraints. Future studies with later ABCD follow-ups will need to be assessed to track growing racial/ethnic disparities in substance use.

## Conclusion

Our findings support national data and extend these findings by showing that ethnic discrimination is associated with greater substance use risk across all racial/ethnic groups in the U.S. Future work should further assess how discrimination and cumulative stress may help to explain this increased risk of substance use. Future research should also assess mediators related to substance use and intentions among each racial/ethnic group. These findings may motivate efforts to address recent racial differences among U.S. adolescents before they transition into adulthood.

## Data Availability

All data produced are available online at https://nda.nih.gov/general-query.html?q=query=featured-datasets:Adolescent%20Brain%20Cognitive%20Development%20Study%20(ABCD)

## Notes

### Competing Interest Statement

The authors have declared no competing interest.

### Funding Statement

This work was supported by the National Institute on Minority Health and Health Disparities (K08MD015289), National Cancer Institute (R01CA270546), and the National Institute on Drug Abuse (R01DA031160; K23DA051561). The content is solely the responsibility of the authors and does not necessarily represent the official views of the National Institutes of Health, the National Institute on Minority Health and Health Disparities, the National Cancer Institute, or the National Institute on Drug Abuse.

### Author Declarations

The study used (or will use) ONLY openly available human data that were originally located at: https://nda.nih.gov/general-query.html?q=query=featured-datasets:Adolescent%20Brain%20Cognitive%20Development%20Study%20(ABCD)

